## Supplemental Table 1 for "Effectiveness of dyadic interventions in improving outcomes for adults with multiple long-term conditions and/or frailty and their informal carers: A systematic review protocol"

### **S1 Table. Search strategy for MEDLINE (Ovid interface)**

1. Multimorbidity/

2. Chronic Disease/

3. Comorbidity/

4. (multimorbid* or multi-morbid* or chronic disease$ or comorbid* or co-morbid* or polymorbid* or poly-morbid* or multidisease* or multi-disease* or disease cluster* or multiple long-term condition* or multiple chronic disease$).tw.

5. ((coocur* or co-ocur* or coexist* or co-exist* or multipl* or concord* or discord*) adj3 (disease$ or ill* or care or condition$ or disorder* or health* or symptom* or syndrom*)).tw.

6. or/1-5
7. Frailty/
8. Frail Elderly/
9. Frailty Syndrome/
10. (frail* or frail* syndrome or geriatric* syndrom* or vulnerabil* or function*).tw.
11. or/7-10
12. 6 or 11

13. Adult/

14. Aged/

15. (adult* or old* or elder* or geriatric* or gerontol* or ageing or aged).tw.

16. or/13-15
17. Spouses/ or Sexual Partners/ or Family/ or Caregivers/
18. (dyad* or pair* or couple* or spouse* or marri* or husband* or wife or wives or romantic partner* or family or families or relative* or friend*).tw.
19. (informal carer* or informal caregiver* or unpaid carer* or unpaid caregiver* or family carer* or family caregiver* or care partner* or support person*).tw.
20. or/17-19
21. ((dyad* or dyadic or pair* or pair-wise or pair-based or couple* or couple-based or two person*) adj3 (intervention* or approach* or coping or therap* or program* or strategy or initiative*)).tw.
22. 12 and 16 and 20 and 21
23. Review/
24. Comment/
25. Letter/
26. Editorial/
27. or/23-26
28. 22 not 27
29. limit 28 to yr="2010 -Current"
